# Feeling the heat: Investigating interoception and motivation as risk factors for exertional heatstroke

**DOI:** 10.1101/2022.08.12.22278401

**Authors:** Charles Verdonk, Camille Mellier, Keyne Charlot, Arnaud Jouvion, Marion Trousselard, Emmanuel Sagui, Alexandra Malgoyre, Pierre-Emmanuel Tardo-Dino

## Abstract

**Background:** Exertional heatstroke (EHS) is the most severe form of heat-related illness, occurring during sport competition or military training. Despite substantial progress in understanding its physiological mechanisms, current evidence suggests the need for broader models that also consider cognitive factors.

**Methods:** We propose a cognitive model of EHS and conduct a preliminary empirical validation through a case-control study using self-report measures. The central hypothesis is that EHS results from a disrupted cost-benefit trade-off during prolonged physical activity, specifically, an overvaluation of performance-related benefits due to excessive motivation, coupled with an undervaluation of exertion costs linked to low interoceptive awareness, characterized by disrupted processing of signals related to the body’s internal state.

**Results:** Individuals with a history of EHS (cases, N=51) reported significantly lower interoceptive awareness and reduced trait mindfulness compared to controls (n=43). However, no difference was found in global motivation traits between groups.

**Conclusion:** These findings provide initial support for a cognitive model of EHS and suggest that simple self-report tools may help identify individual vulnerability. Incorporating cognitive dimensions into EHS research could enhance risk stratification and inform new prevention strategies in athletic and military contexts.

## INTRODUCTION

Exertional heatstroke (EHS) is the most serious condition in the spectrum of heat illnesses that can occur during sport competition or physical activity within specific contexts such as military training (Epstein and Yanovich 2019). The incidence of EHS remains relatively low in sport competitions (Stearns, Hosokawa et al. 2017), however it might become a major health concern in the future because of global warming. Previous research has revealed a large number of risk factors for EHS, including extrinsic factors (e.g., environmental stress such as high temperature or high humidity) as well as intrinsic factors (i.e., individual-specific factors, such as sleep deprivation or alcohol consumption) (Gardner, Kark et al. 1996, Abriat, Brosset et al. 2014, Epstein and Yanovich 2019). From a physiological standpoint, EHS is classically described as a non-compensable heat stress where heat loss does not balance heat gain during a prolonged physical effort (see the Supplementary Introduction for a graphical overview of the suspected physiological mechanisms of EHS) (Epstein and Yanovich 2019, Laitano, Leon et al. 2019). Despite substantial progress in understanding pathophysiology of EHS, which has contributed to the development of preventive strategies for reducing the risk of EHS in sport competitions (Parsons, Anderson et al. 2020, Mountjoy, Moran et al. 2021), the current models are still limited to inform the risk of EHS at the individual level.

Interestingly, several studies have highlighted that cognitive factors (e.g., motivation) can influence the injury risk during sport competition (Ivarsson, Johnson et al. 2017). For instance, overmotivation has been suggested as a potential risk factor for EHS on the basis of investigation of a relatively small cohort of fatal cases (Rav-Acha, Hadad et al. 2004). More recently, Corbett et al. (2017) have investigated the impact of motivation on thermophysiological strain, and they have demonstrated that competition-induced overmotivation leads to increased thermophysiological cost that may not be perceived (consciously) by the participant (Corbett, White et al. 2017). This finding suggests that overvaluation of benefits, resulting from overmotivation to succeed, might contribute to the disruption of cost-benefit trade-off that characterizes adjusted physical effort. The equilibrium between costs and benefits is thought to be a central determinant of behavior. In elite sport, every workload decision - whether made by a coach or an athlete - reflects a trade-off between performance gains and injury risk. Gabbett et al. (2016) exemplify this using a dynamic metric (the “acute:chronic workload ratio”) that combines short-term performance benefits, accumulated fatigue, and individual variability to optimize training while minimizing harm (Gabbett, Windt et al. 2016).

In the present paper, we introduce a model that characterizes EHS by the alteration of cost-benefit trade-off within the context of prolonged physical effort. Specifically, we propose that EHS could be the consequence of *overvaluation* of benefits in combination with *undervaluation* of costs associated with physical effort (Fig. 1).

**Fig. 1.**
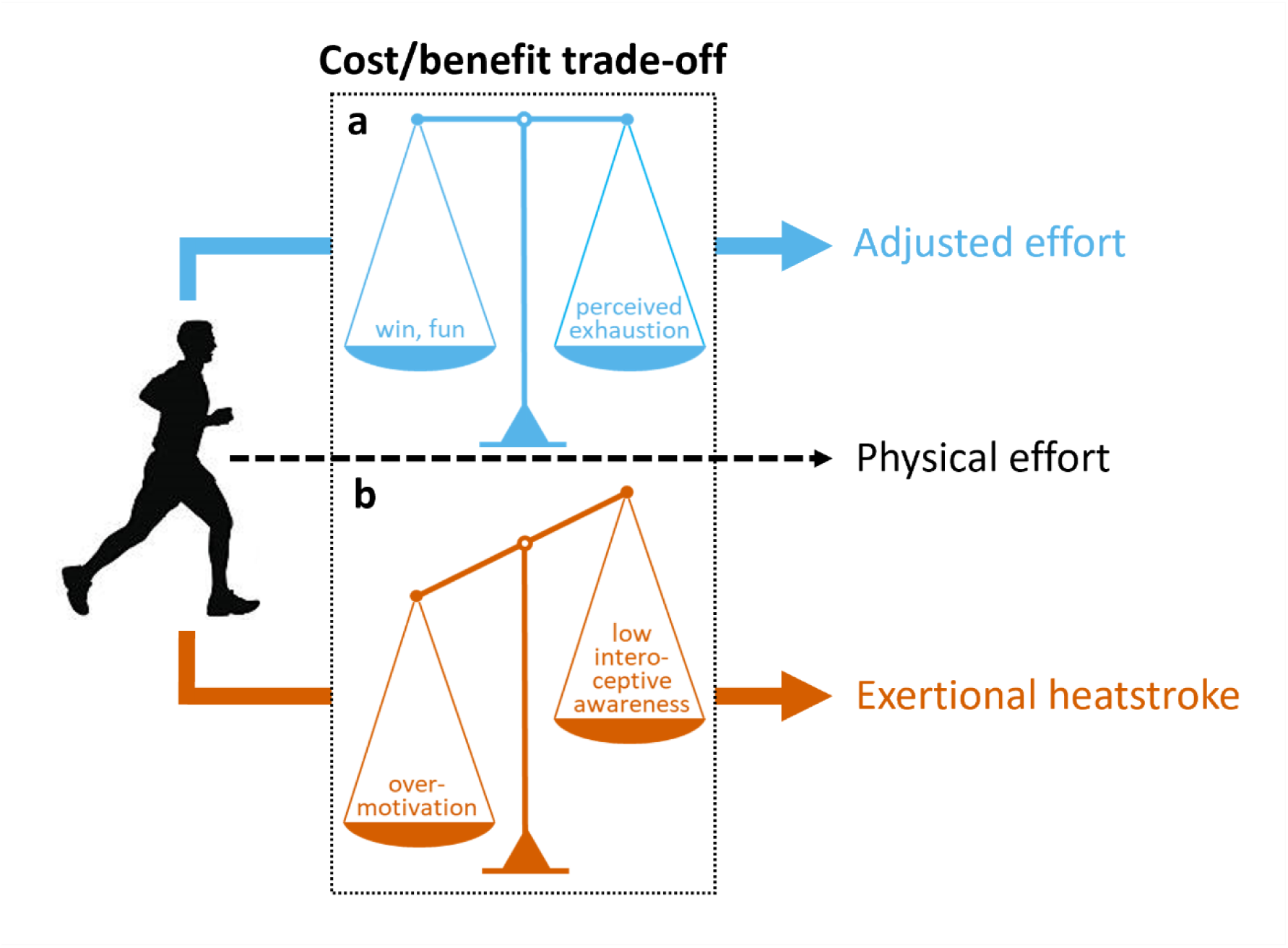
Schematic illustration of the proposed cognitive model for exertional heatstroke. **(a)** Physical effort in safety conditions relies on a sustained balance between the costs and benefits of effort exertion. **(b)** We argue that exertional heatstroke is characterized by alteration of cost/benefit trade-off. Specifically, the overvaluation of benefits associated with physical effort, as a consequence of overmotivation to succeed, and/or the undervalued costs of effort exertion that result from low body awareness, disrupt the cost/benefit trade-off and lead individuals to maintain physical effort, which can ultimately lead to exertional heatstroke.

Our first assumption is that overmotivation might cause individuals to overvalue benefits that they associate with physical effort (e.g., personal satisfaction, win in the context of competition).

Overmotivation has previously been suggested as a potential risk factor for EHS (Rav-Acha, Hadad et al. 2004, Epstein and Yanovich 2019), but it has never been tested empirically. Classical theories of motivation assume that people initiate and persist at behaviors to the extent that they believe the behaviors will meet their needs. In the field of psychology, the term ‘needs’ refers to innate psychological ‘nutriments’ that are essential for ongoing psychological growth, integrity, and well-being. In the self-determination theory of motivation (SDT), three psychological needs— for competence, relatedness, and autonomy - are considered essential for understanding motivated behavior (Deci and Ryan 2000, Ryan and Patrick 2009). SDT distinguishes *intrinsic* and *extrinsic* types of motivation regulating one’s behavior. Briefly, the *intrinsic* motivation is defined as doing an activity because of its inherent satisfaction, e.g., for the enjoyment associated with the activity. By contrast, the *extrinsic* motivation refers to doing an activity for obtaining some outcome separable from the activity *per se*, e.g., for the gain of social reward resulting from win in sport competition. Of note, the SDT conceptualizes qualitatively different types of extrinsic motivation as measured with the Global Motivation Scale (see the Method for detailed description of the scale). In our model, the motivation is considered as a stable psychological trait rooted in the individual’s personality (Scheffer and Heckhausen 2018), i.e. a global motivational orientation, which differs from situational motivation that refers to motivation toward a given activity at a specific point in time, according to the hierarchical model of self-determined motivation (Vallerand 2007).

Our second assumption is that sensitivity to bodily signals might influence how individuals self-regulate their physical activity. Indeed, one can bring awareness to internal body changes caused by physical effort (e.g., heart rate increase, fatigue, hyperthermia) and ascertain the embodied cost of physical activity. If the cost is assessed as being high, the update of the cost-benefit trade-off may lead to a decrease in commitment to physical activity, or even a complete stop, to maintain good safety conditions. By contrast, individuals exhibiting low sensitivity to bodily signals may maintain physical effort despite physiological cost, which can ultimately lead to exertional heatstroke. The cognitive construct of body awareness refers to individuals’ ability to feel engaged by information coming from within their body and noticing subtle changes (Mehling, Gopisetty et al. 2009). From a neurophysiological standpoint, bodily signals continuously provide the brain with a moment by moment mapping of the body’s physiological state, also called interoception, whose integration at higher-order cortical regions, notably the insula, results in the emergence of interoceptive awareness (Craig 2002, Berntson and Khalsa 2021). Interestingly, the insula has been shown to encode cost information during a force task and triggers participants’ decision to stop their physical effort (Meyniel, Sergent et al. 2013). The authors also found that motivation may impact neural processes underpinning effort allocation, specifically by pushing back limits and allowing the body to work closer from exhaustion

Regarding the potential long-term clinical applications of our model, we believe that shedding new light on cognitive factors that influence the risk of EHS could ultimately lead to novel preventive interventions. For instance, if low interoceptive awareness characterizes individuals at high risk for EHS, a relevant prevention strategy could be an intervention that enhances interoceptive awareness via effects on the cognitive processing of bodily signals. Contemplative practice, such as mindfulness meditation, has been shown to enhance body awareness by training the mind to pay sustained attention to the body experience, and deliberately returning attention to it whenever distracted (Lutz, Jha et al. 2015, Treves, Tello et al. 2019). It could be argued that the more fully individuals are apprised of what is occurring within their body, the more adaptive and value-consistent their behaviours are likely to be during physical effort.

To summarize, our cognitive model of EHS suggests that overvaluation of benefits, as a consequence of overmotivation, and undervaluation of costs associated with physical activity, resulting from low interoceptive awareness, could lead individuals to ignore body warning signs of EHS (e.g., hyperthermia, tachycardia, tachypnea) thus preventing any attempt to self-regulate physical activity in safety conditions (Fig. 1). In the present study, we confronted our model to psychometric data including self-reported trait motivation, interoceptive awareness, and trait mindfulness, in a cohort of subjects with or without a history of EHS. We predicted that subjects with a history of EHS (cases) would show higher levels of trait motivation, lower interoceptive awareness, and lower levels of trait mindfulness, compared to healthy subjects (controls).

## METHODS

### 1. Participants

We recruited 51 patients with a history of EHS (cases) from the French Military Teaching Hospital Laveran (Marseille, France), between 2014 and 2020. Cases were service personnel from conventional combat units of the French Army who have experienced EHS during training or operation, and whose diagnosis and follow-up care were managed by medical teams from the French Military Health Service. Cases completed self-report questionnaires during their routine follow-up visits at the hospital. Data for the present analysis were collected 1244 ± 199 days (mean ±SEM) after the EHS event. There was no exclusion criterion for cases. However, all cases were active-duty service personnel who regularly underwent medical evaluations to ensure operational readiness, thus limiting significant health comorbidities.

Controls (n=43) were recruited between March 9, 2021 and May 12, 2021, from several French Army units involved in territorial surveillance duties. Like the cases, control participants were career soldiers from conventional operational. To ensure comparable exertional exposure, all controls had, at some point during their service, completed an 8 km run in battledress - a standard exercise that challenges thermal balance and during which most EHS cases occur in the French Army (Abriat, Brosset et al. 2014). Controls were matched to cases by morphological characteristics (height and weight), sex and age. All controls reported no history of exertional heatstroke, no ongoing medication, and no past or present psychiatric or somatic disorders. Additionally, to ensure relevant operational experience, only soldiers with at least 18 months of service and prior exposure to physical fitness testing and/or operational missions in hot climates were included. Controls completed the self-report questionnaires in a dedicated experimental session that was planned during their spare time. This study was approved by the regional ethics committee of the Agence Régionale de Santé Occitanie (Comité de Protection des Personnes Sud-Ouest et Outre-Mer II, ID-RCB: 2020-A01967-32) on September 29, 2020. Written informed consent was obtained from all individual participants included in the study. The study was conducted in accordance with ethical standards of the 1964 Helsinki declaration and its later amendments.

## 2. Cognitive self-reported measures

### 2.1. *Interoceptive body awareness*

The 32-item Multidimensional Assessment of Interoceptive Awareness (MAIA) questionnaire measures eight facets of body awareness: (1) *Noticing*: awareness of uncomfortable, comfortable, and neutral body sensations; (2) *Not-distracting*: tendency not to be distracted by oneself from sensations of pain or discomfort; (3) *Not-worrying*: tendency not to worry with sensations of pain or discomfort; (4) *Attention regulation*: ability to sustain and control attention to body sensation; (5) *Emotional Awareness*: awareness of the connection between body sensations and emotional states; (6) *Self-regulation*: ability to regulate psychological distress by attention to body sensations, (7) *Body listening*: actively listens to the body for insight, and (8) *Trusting*: experiences one own’s body as safe and trustworthy. The questionnaire is scored using a six-point scale, with responses ranging from 0 (never) to 5 (always). For each of the eight subscales, the score was counted by averaging the scores of items belonging to each subscale (items 5, 6, 7, 8 and 9 were reversed) (Mehling, Price et al. 2012, Willem, Gandolphe et al. 2021). In the present work, the MAIA questionnaire demonstrated acceptable levels of internal consistency in Cases (Cronbach alpha = 0.90) and in Controls (Cronbach alpha = 0.89).

### 2.2. Self-determined motivation

The 28-item Global Motivational Scale (GMS) assesses three types of intrinsic motivation (IM): (1) *IM to knowledge* : pleasure while learning, exploring or trying to understand something new, (2) *IM* t*o accomplishment* : pleasure to accomplish or create something, and (3) *IM to stimulation* : pleasure to have a stimulating discussion or intense feelings of cognitive pleasure; three types of extrinsic motivation (4) *identified regulation* : doing something because it matches ones values, *introjected regulation* : doing something because it is supposed to be good for oneself, and *external regulation* : doing something in order to have a reward or to avoid punishment ; and *amotivation* : lack of extrinsic or intrinsic motivation. The questionnaire is scored using a seven-point scale with responses ranging from 1 (not at all) to 7 (totally). For each of the seven subscales assessed by 4 items, the score was counted by averaging the scores of items belonging to each subscale (Vallerand, Pelletier et al. 1992). In the present work, the GMS demonstrated acceptable levels of internal consistency in Cases (Cronbach alpha = 0.88) and in Controls (Cronbach alpha = 0.93).

### 2.3. Trait mindfulness

The 14-item Freiburg Mindfulness Inventory (FMI) measures dispositional trait mindfulness by indexing facets of Presence (*i.e.* being aware of all experiences in the present moment) and Non-judgmental acceptance (*i.e.* understanding that things are not necessarily how one wishes them to be). This questionnaire is semantically independent from a meditation context and it is applicable to all population groups, in particular to those with no practice of mindfulness meditation. The questionnaire is scored using a four-point scale, with responses ranging from 1 (rarely) to 4 (almost always). A total mindfulness score was computed by adding the rating for all items, except for the 13th item which was reversely scored (Walach, Buchheld et al. 2006, Trousselard, Steiler et al. 2010). In the present work, the FMI demonstrated acceptable levels of internal consistency in Cases (Cronbach alpha = 0.80) and in Controls (Cronbach alpha = 0.77).

### 3. Statistical analyses

Data analyses were performed using JASP (version 0.11.1, https://jasp-stats.org/). We used both standard statistical tests and Bayesian equivalents to extend insight and guiding interpretation of significance (p values), according to how likely the alternative hypothesis is versus the null. Indeed, a disadvantage of null hypothesis significance testing is that non-significant p values (e.g., when reporting no significant difference between Cases and Controls for self-reported trait motivation) cannot be interpreted as support for the null hypothesis (Rouder, Speckman et al. 2009, Wagenmakers, Marsman et al. 2018). To circumvent this issue and confirm whether the potential non-significant findings reported represent support for the null hypothesis, we calculated the Bayes factor (BF): specifically, we computed the log scale of BF_10_ (noted log(BF_10_)) that can be easily interpreted such that a negative value indicates support for the null hypothesis, whereas a positive value indicates evidence in favour of the alternative hypothesis (see *Supplementary Table 1* for an interpretation scale of log(BF_10_) (Jeffreys 1961)). Statistical analyses were performed using Mann-Whitney nonparametric tests, as data from Cases and Controls were not normally distributed. If a significant difference was observed, we computed the effect size (to evaluate the magnitude of the difference) using a measure suited to nonparametric analyses: 95% confidence interval of the rank biserial correlation (Glass 1966). For the Bayesian analyses, we used the default JASP priors that assume a medium effect size on a Cauchy distribution of 0.707 for independent t-tests.

As this is the first study to explore interoception and motivation via self-report measures in an EHS cohort, no prior data were available to inform an a priori power or sample-size calculation. However, to ensure that our analyses were nevertheless adequately powered, we conducted post hoc power analyses on significant data (see the Supplementary Methods).

## RESULTS

### 1. Demographic and biometric characteristics

Table 1 summarizes the basic statistics on demographic (age and gender) and biometric (weight, height, and body mass index) measures in cases and controls. Cases and controls did not differ for age (log(BF_10_) = -1.37, suggesting strong evidence for the null hypothesis), gender (log(BF_10_) = -2.03, suggesting extreme evidence for the null hypothesis), weight (log(BF_10_) = - 1.50, suggesting very strong evidence for the null hypothesis), height (log(BF_10_) = -1.43, suggesting strong evidence for the null hypothesis), and body mass index (log(BF_10_) = -1.44, suggesting strong evidence for the null hypothesis).

**Table 1:** Summary of demographic (age and gender) and biometric (weight, height, and body mass index) data for cases with a history of exertional heatstroke (cases) and controls.

|  | **Cases**<br>(n=51) | **Controls**<br>(n=43) | P-value<sup>†</sup> | log(BF<sub>10</sub>)<sup>†</sup> |
| --- | --- | --- | --- | --- |
| *Characteristics* |  |  |  |  |
| **Age** (years), M (SD) | 27.80 (6.38) | 27.30 (6.48) | 0.65 | -1.37 |
| **Male gender**, n (%) | 48 (94) | 40 (93) | 0.83 | -2.03 |
| **Weight** (kg), M (SD) | 76.94 (8.41) | 77.28 (9.33) | 0.89 | -1.50 |
| **Height** (cm), M (SD) | 176.24 (7.80) | 177.02 (5.17) | 0.67 | -1.43 |
| **Body Mass Index**<br>(kg.cm<sup>-2</sup>), M (SD) | 24.79 (2.45) | 24.65 (2.76) | 0.57 | -1.44 |

<sup>†</sup> Mann-Whitney nonparametric tests (continuous data) and $\chi^2$ test (categorical data); M: mean; SD: standard deviation; kg: kilograms; cm: centimeters; log(BF<sub>10</sub>): log scale of Bayes factor BF10

### 2. Interoceptive body awareness

Five dimensions of interoceptive body awareness were significantly lower in cases compared to controls: *Body listening* (Mann-Whitney U (U) = 717, p ≤ 0.01, 95% confidence interval (CI) of rank-biserial correlation (rbs) = [0.12 - 0.54]), *Attention regulation* (U = 730, p ≤ 0.01, 95% CI of rbs = [0.11 - 0.53]), *Emotional awareness* (U = 767, p ≤ 0.05, 95% CI of rbs = [0.07 - 0.50]), *Self-regulation* (U = 780, p ≤ 0.05, 95% CI of rbs = [0.06 - 0.49]), and *Noticing* (U = 839, p ≤ 0.05, 95% CI rbs = [0.003 - 0.44]). Furthermore, cases and controls did not differ for three dimensions of interoceptive body awareness: *Not-worrying* (log(BF_10_) = -1.36, suggesting strong evidence for the null hypothesis), *Trusting* (log(BF_10_) = -0.39, suggesting moderateevidence for the null hypothesis), and *Not-distracting* (log(BF_10_) = -0.20, suggesting anecdotal evidence for the null hypothesis). Fig. 2 summarizes how cases and controls differ in terms of self-reported interoceptive body awareness. Descriptive statistics of self-reported interoceptive body awareness in cases and controls are reported in *Supplementary Table 2*. All significant dimensions of interoception (Body listening, Attention regulation, Emotional awareness, Self-regulation, and Noticing) produced effect sizes in the medium-to-large range, with achieved power between 0.79 and 0.94 (see *Supplementary Table 5*), indicating sufficient sensitivity to detect the reported group differences.

**Fig. 2.**
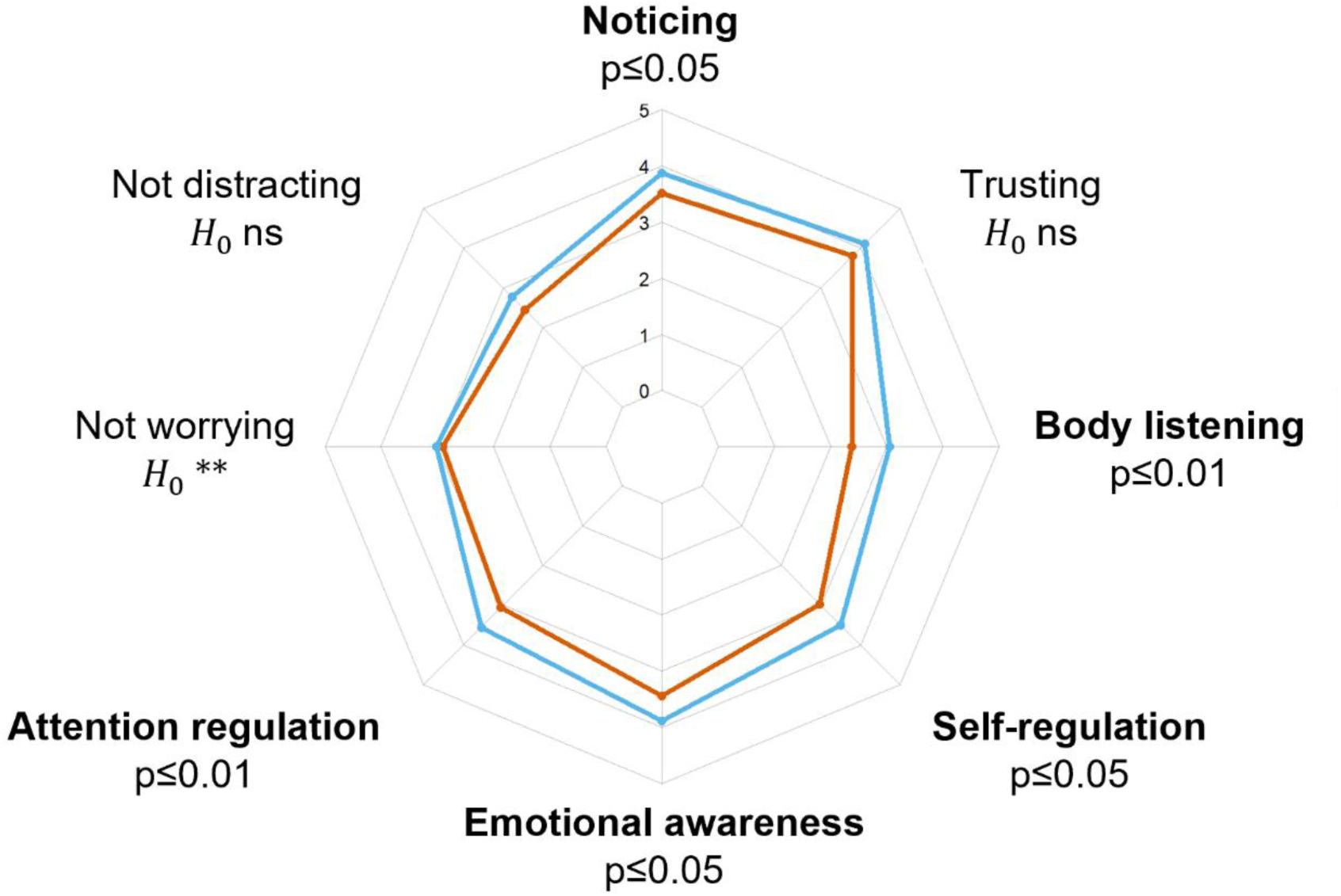
Graphical comparison of cases with a history of exertional heatstroke (orange line) and controls (blue line) with respect to eight facets of interoceptive body awareness, as assessed with the Multidimensional Assessment of Interoceptive Awareness questionnaire. Significant (p<0.05) differences are highlighted in bold. *Note :* 𝐻_0_*ns: anecdotal evidence for the null hypothesis;* 𝐻_0_*\*\*: strong evidence for the null hypothesis*.

### 3. Self-determined motivation

Cases and controls did not differ for almost all factors of motivation: *IM to accomplishment* (log(BF_10_) = -1.50, suggesting very strong evidence for the null hypothesis), *Introjected regulation* (log(BF_10_) = -1.53, suggesting very strong evidence for the null hypothesis), *Identified regulation* (log(BF_10_) = -1.53, suggesting very strong evidence for the null hypothesis), *IM to stimulation* (log(BF_10_) = -1.51, suggesting very strong evidence for the null hypothesis), *IM to know* (log(BF_10_) = -1.21, suggesting strong evidence for the null hypothesis), and *External regulation* (log(BF_10_) = -0.90, suggesting moderate evidence for the null hypothesis). Only the factor *Amotivation* was significantly lower in cases compared to controls (U = 729, p ≤ 0.01, 95% CI of rbs = [0.11 - 0.53]). Fig. 3 summarizes similarities between cases and controls in terms of self-determined motivation. Descriptive statistics of self-determined motivation in cases and controls are reported in *Supplementary Table 3*. The motivation dimension “Amotivation” demonstrated a large effect size and achieved power of 0.93 (see *Supplementary Table 5*), confirming sufficient sensitivity to detect the observed group difference.

**Fig. 3.**
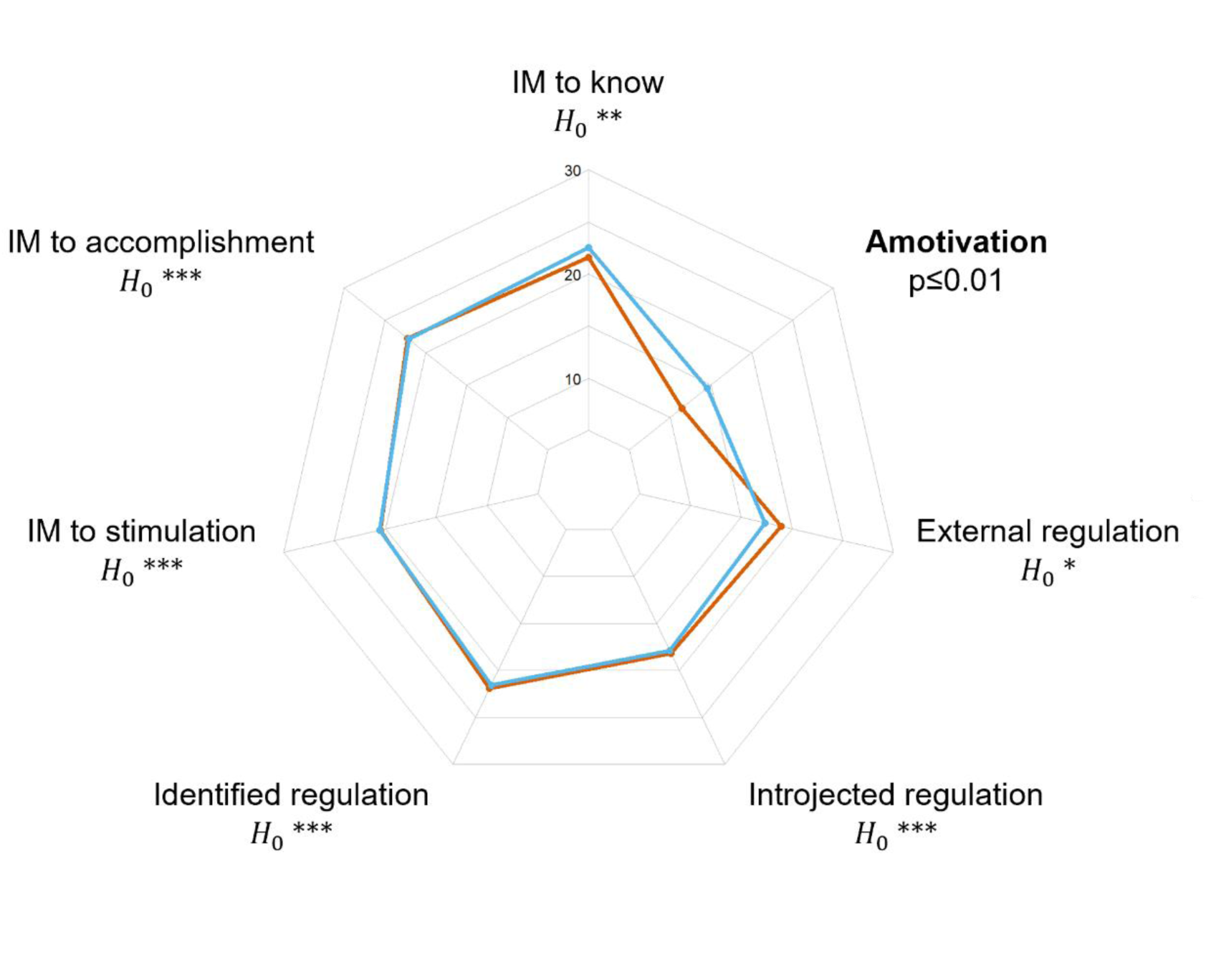
-. Graphical comparison of cases with a history of exertional heatstroke (orange line) and controls (blue line) with respect to seven factors of self-determined motivation, as assessed with the Global Motivation Scale. The only significant difference is highlighted in bold. *Note : \** 𝐻_0_*\*: moderate evidence for the null hypothesis;* 𝐻_0_*\*\*: strong evidence for the null hypothesis*; 𝐻_0_*\*\*\*: very strong evidence for the null hypothesis*.

### 4. Trait Mindfulness

Cases showed lower scores for the two mindfulness dimensions that are assessed with the FMI, relative to controls: *Presence* (U = 761, p ≤ 0.05, 95% CI of rbs = [0.08 - 0.50]), and *Acceptation* (U = 730, p ≤ 0.01, 95% CI of rbs = [0.11 - 0.52]) (Fig. 4). Descriptive statistics of trait mindfulness in cases and controls are reported in *Supplementary Table 4*.

**Fig. 4.**
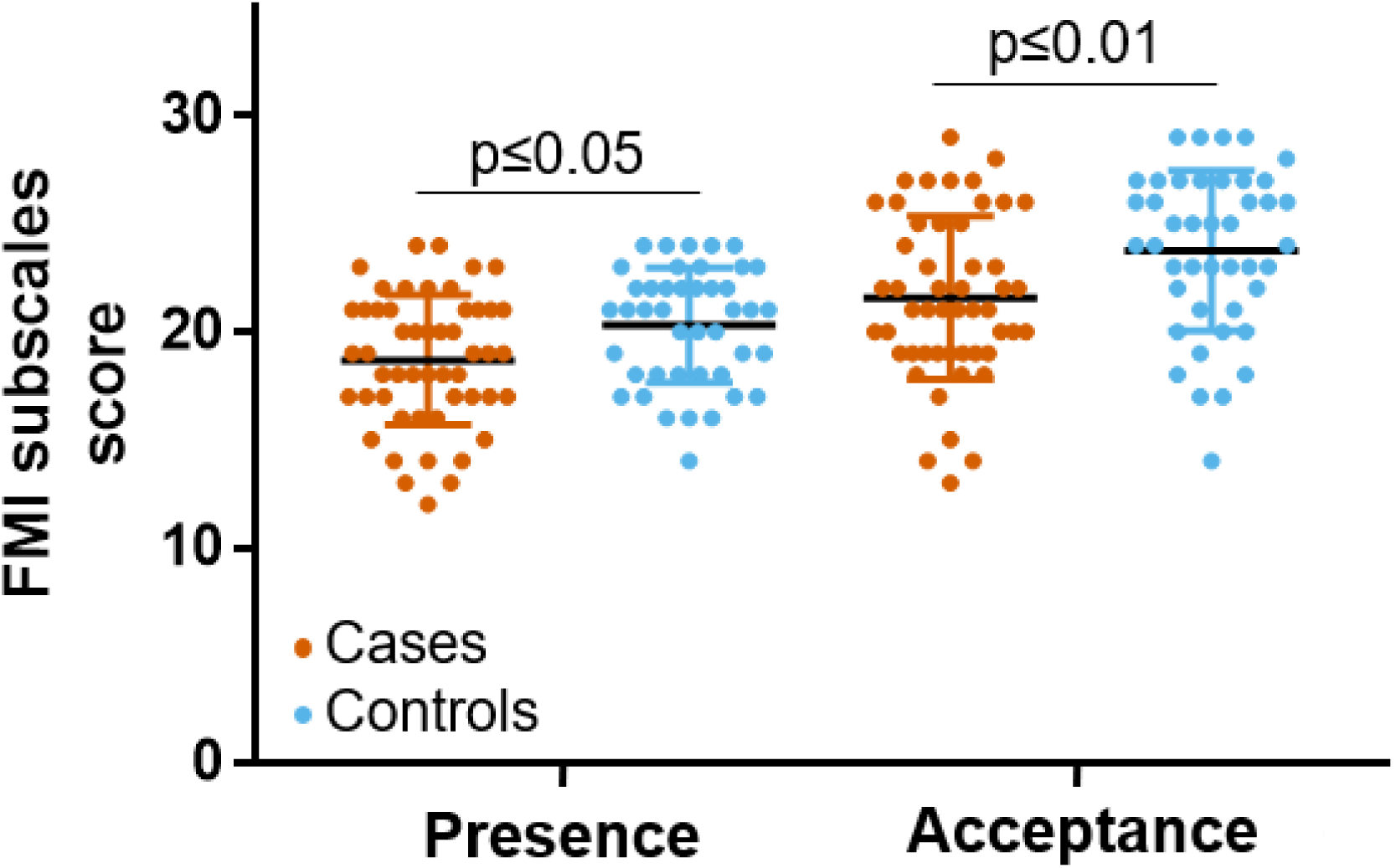
Significant differences were found between cases with a history of exertional heatstroke (in orange) and controls (in blue) for mindfulness dimensions of *Presence* and *Acceptance*, as assessed with the Freiburg Mindfulness Inventory (FMI).

## DISCUSSION

### General discussion

This paper introduces a testable model that explores cognitive factors influencing the risk of exertional heatstroke (EHS). Our theoretical framework encompasses two key components: the self-determination theory of motivation (Deci and Ryan 2000, Ryan and Patrick 2009) and the concept of body awareness (Mehling, Gopisetty et al. 2009). At its core, our hypothesis posits that EHS may arise from a disruption in the cost-benefit trade-off associated with prolonged physical activity (Fig. 1). Specifically, we suggest that EHS could be the result of an *overvaluation* of the benefits linked to physical activity due to excessive motivation to succeed and, simultaneously, there may be an *undervaluation* of the physiological costs associated with effort exertion, stemming from low interoceptive awareness, i.e. disrupted processing of signals about the body’s internal state. To test our theoretical model, we conducted a study in a cohort of subjects, distinguishing between those with a history of exertional heatstroke (cases) and those without (controls), utilizing self-report measures of motivation and interoceptive awareness. Although questionnaires were administered on average 1244 days after the EHS event (range: 800–1800 days), this long interval likely minimizes any acute vigilance or trauma-related bias in interoceptive self-reports; a shorter delay might have captured transient, event-driven states rather than more stable, trait-like differences.

The primary observation reveals that cases reported significantly lower interoceptive awareness compared to controls across the dimensions of Body listening, Attention regulation, Emotional awareness, Self-regulation, and Noticing, as assessed through the MAIA questionnaire (Mehling, Price et al. 2012, Willem, Gandolphe et al. 2021). The diminished scores on the Body Listening, Attention Regulation, and Self-Regulation subscales suggest that individuals with a history of EHS may encounter challenges actively and consistently listening to their body to gain ongoing insights into their body’s internal state. In the context of physical activity, this could signify a potential difficulty in detecting physiological signs of exhaustion (e.g., hyperthermia, tachycardia, tachypnea) that may prevent individuals from self-regulating their physical effort, thus increasing the risk of EHS. Moreover, the low score on the Noticing subscale indicates that individuals with a history of EHS, while potentially able to sense some body signals, struggle to differentiate between negative body sensations (e.g., warning body signs of EHS) and sensations associated with physical effort in safe conditions. The reduced score on the Emotional Awareness subscale, reflecting difficulty attributing specific physical sensations to physiological manifestations of emotions (Mehling, Price et al. 2012, Willem, Gandolphe et al. 2021), suggests that individuals with a history of EHS may be less responsive to negative stimuli both within their bodies and in the external environment. Taken together, these findings may pave the way for the development of novel prevention strategies aimed at reducing the risk of EHS. Specifically, it is conceivable that interventions focusing on enhancing the cognitive process of interoceptive awareness could play a crucial role in mitigating the risk of EHS. By training individuals to better perceive and interpret body sensations, such programs have the potential to foster heightened interoceptive awareness, which in turn may empower individuals to implement more effective self-regulation strategies during physical activity, thereby reducing the likelihood of EHS occurrences. Moreover, targeted health-education messages for military personnel, athletes, and the general public can be developed from these preliminary findings. Such messages should emphasize improving active attention to one’s bodily signals and learning individualized warning signs. Elite athletes - who often cultivate advanced body-listening skills through high-level training and mental rehearsal - illustrate how this capacity can be strengthened (Wallman-Jones, Perakakis et al. 2021). Translating these athlete-tailored strategies to broader populations could significantly bolster prevention efforts for EHS.

The second main observation is that cases demonstrated a less developed trait mindfulness compared to controls, as assessed with the FMI (Walach, Buchheld et al. 2006, Trousselard, Steiler et al. 2010). It is pertinent to note that engaging in mindfulness practice has been demonstrated to foster the development of trait mindfulness (Brown and Ryan 2003, Kiken, Garland et al. 2015) and enhance body awareness by training the mind to pay attention to body sensations (e.g., through the body scan exercise) (Treves, Tello et al. 2019). These findings suggest that a mindfulness-based program, designed to train individuals to access their body sensations deliberately and continuously, may be a promising candidate as a prevention strategy to mitigate the risk of EHS. It is important to emphasize that only studies manipulating mindfulness (e.g., through mindfulness intervention) will offer conclusive evidence regarding whether mindfulness practice causally reduces the risk of EHS. We recognize that conducting such studies poses a significant methodological challenge because of the perceived rarity of EHS events (Stearns, Hosokawa et al. 2017, Epstein and Yanovich 2019).

The final observation in our study reveals that individuals with a history of EHS did not exhibit differences from healthy subjects concerning motivational trait (i.e., global motivation), as assessed by the GMS (Vallerand, Pelletier et al. 1992). Notably, the use of Bayesian statistical tests allowed us to provide moderate to strong evidence in favor of the absence of difference between cases and controls, rather than a simple declaration that the null hypothesis cannot be statistically rejected (Rouder, Speckman et al. 2009, Wagenmakers, Marsman et al. 2018).

Although this finding is confined to self-report global motivation measure (as discussed in our limitations section), it contributes to a psychometric argument derived from the GMS against a key assumption in our theoretical model, specifically that EHS could arise from an overmotivation to succeed (Fig. 1). At present, it remains uncertain whether the lack of association observed between global motivation and history of EHS is pertinent to refine our model in suggesting that the factor motivation may be not relevant to inform the risk of EHS, or whether it reflects intrinsic limitations of self-report measures (e.g., limitations of introspection, social-desirability biases; (Baumeister et al., 2007)) to capture the cognitive construct of motivation.

In addition, our model suggest the relevance of a broader construct of “body-related self-regulatory competence”, which integrates three key elements: (1) the ability to accurately sense internal signals (interoceptive awareness), (2) the capacity to interpret and respond to those signals through appropriate self-regulation strategies, and (3) the health-literacy skills needed to understand the significance of bodily warning signs (Fazekas, Avian et al. 2022, Fazekas, Linder et al. 2022). Our current findings document differences only at the first level - accurate sensing of internal signals (interoceptive awareness). However, dysregulation at subsequent levels - namely, interpreting bodily cues correctly and enacting appropriate self-regulation - may also contribute to EHS risk. Although the MAIA captures multiple facets of interoceptive awareness (including a “Self-Regulation” subscale; (Mehling, Price et al. 2012, Willem, Gandolphe et al. 2021)), it does not assess behavioral adjustment or health-literacy skills. Future research should therefore employ ecological momentary assessment of effort adjustments and performance-based paradigms that directly link signal detection to behavioral change, alongside instruments assessing health literacy, to test these additional regulatory levels and their cumulative impact on EHS susceptibility.

Moreover, EHS can also result from acute impairments of consciousness that bypass any cost–benefit evaluation. Heat-induced dizziness, confusion, or cognitive slowing can immediately degrade one’s ability to perceive and interpret interoceptive signals, as well as to enact self-regulatory behaviors and sound decision-making. While our current self-report measures at rest reveal baseline differences between cases and controls, we anticipate these impairments would become even more pronounced under heat or exertion conditions. In such situations, individuals - even those with intact trait motivation and self-regulatory competence - may be unable to recognize or respond to warning signs in real time. We therefore propose that future studies integrate both resting and real-time assessments of cognitive function and conscious state (e.g., brief cognitive tasks during exertion) alongside trait measures, to capture the full spectrum of chronic vulnerabilities and acute cognitive disruptions that contribute to EHS.

### Limits of the theoretical model and its preliminary testing

For simplicity, our cognitive model of EHS only considers the trait component of motivation that refers to the global motivational orientation of individuals at the personality level. According to the hierarchical model proposed by Vallerand (2007), self-determined motivation can be described at additional levels of generality. The lowest level of generality corresponds to the situational motivation, which pertains to the motivation experienced by an individual toward a given activity at a specific point in time (Vallerand 2007). In our work, we did not measure the situational motivation because psychometric data were collected outside any context of physical activity. Our data show that self-reported global motivation (trait motivation) does not help differentiate subjects with a history of EHS from healthy subjects. This suggests that limiting assessment of the motivational factor to its last level of generality (*i.e.* the personality level) might not be informative about the risk of EHS. Future studies are encouraged to investigate the situational motivation as a potential alternative factor that could influence the risk of EHS. It could be suggested that normal (or low) global motivation combined with a high situational motivation could ultimately result in a high level of self-determined motivation. In other words, motivational factors from different levels of generality could potentially have a cumulative effect on how individuals are engaged in an activity. To test this hypothesis, experimental settings need to include measurements of both trait motivation and situational motivation, or experimentally manipulate situational motivation of the participant by using incentive motivation paradigm (Pessiglione, Schmidt et al. 2007). Classically, measurement of situational motivation relies on self-report instruments, such as the Situational Motivation Scale (Guay, Vallerand et al. 2000, Clancy, Herring et al. 2017). Yet, self-report instruments are often criticized because they may be vulnerable to limitations of introspection and social-desirability biases, and are potentially limited by individual’s unwillingness or inability to report their veridical cognitive state (Baumeister et al., 2007). We argue that even self-report instruments provide valuable information and are particularly attractive for field research, they should not be considered in isolation in future cognitive research into EHS. Interestingly, some works combining behavioural measures and neurocomputational models of motivation have opened promising opportunities to address the aforementioned issue related to self-report questionnaires. For instance, in the incentive motivation task developed by Pessiglione *et al* (2007), behavioral measures (e.g. the peak of force with which the participant squeezes the power grip) can be modeled as functions that approximate the solutions of an optimal motor-control model (which maximizes the cost/benefit tradeoff) at the individual level (Pessiglione, Schmidt et al. 2007, Le Bouc, Rigoux et al. 2016).

Such a neurocomputational approach has the advantage of providing motivation related measures (e.g. the parameter of expected reward) that are not contaminated by individual differences in other cognitive components (e.g., emotional thoughts, changes in attention, etc.). Therefore, we encourage future studies investigating motivational underpinnings of EHS to use this neurocomputational approach that has great potential to enhance the process of relating differences (in behaviour and neural processes) between healthy subjects and individuals who are at risk of EHS.

Regarding the other main component of our model that refers to interoceptive body awareness, it has been formalized as a multifaceted cognitive process that can be interrogated with complementary methods (Garfinkel, Seth et al. 2015, Khalsa and Lapidus 2016), including self-report instruments and objective measurements (e.g., behavioural test or biomarker). Objective measurements of interoceptive body awareness mainly focus on cardiac interoception that refers to the process of sensing, storing and representing information about the state of the cardiovascular system (Garfinkel, Seth et al. 2015, Khalsa and Lapidus 2016). These measurements are mostly performed under conditions of physiological rest, *i.e.* without any significant experimentally-induced cardiovascular manipulation, which raises questions about their potential relevance to inform interoceptive dysfunction in the context of EHS. Indeed, physical effort is characterized by strong, continuous perturbations that affect the cardiovascular system (e.g. increased heart rate and arterial pressure). Interestingly, the pharmacological manipulation of cardiac arousal (via the administration of isoproterenol that modulates sympathetic nervous system) may provide an attractive experimental framework, because it has the advantage of a maskable manipulation of arousal (including placebo condition) that allows for measurements of responding bias (Khalsa, Rudrauf et al. 2009). Beside behavioural tests that provide an indirect output of interoceptive signal processing, the neural bases of cardiac interoception can be investigated by probing brain activity in response to cardiac signals. The Heartbeat Evoked Potential, which refers to evoked changes in brain activity (measured using magnetoencephalography, electroencephalography, or intracranial neural recordings) that occurs after a heartbeat, has been proposed as a neurophysiological marker of interoceptive function/dysfunction (Park and Blanke 2019, Coll, Hobson et al. 2021).

To summarize, future studies are encouraged to pursue the development and validation of a cognitive model for EHS in using neurocomputational approach of motivation with the incentive motivation paradigm (Pessiglione, Schmidt et al. 2007), and objective measurements of interoceptive body awareness (e.g., heartbeat perception task (Brener and Ring 2016) or analysis of the heartbeat evoked potential (Park and Blanke 2019, Coll, Hobson et al. 2021)) based on a paradigm involving pharmacological manipulation of cardiac arousal (Khalsa, Rudrauf et al. 2009).

## CONCLUSION

In this study, we proposed and provided preliminary empirical support for a cognitive model of exertional heatstroke (EHS). Individuals with a history of EHS exhibited significantly diminished interoceptive awareness and reduced trait mindfulness compared to controls, but did not differ in global motivation trait. These results identify low interoceptive awareness as a potential intrinsic risk factor for EHS among military personnel and emphasize the importance of assessing interoception to inform return-to-duty decisions. Our findings encourage future research on mindfulness-based preventive strategies and the assessment of situational motivation as additional pathways for reducing EHS risk in athletic and military contexts.

## Data Availability

The data and materials are currently private for peer review.

## Author Notes

This study is part of a project supported by the French Military Health Service. The opinions or assertions expressed herein are the private views of the authors and are not to be considered as official or as reflecting the views of the French Military Health Service.

## Acknowledgments

We thank Walid Bouaziz, Caroline Dussaud, Julien Siracusa, Stéphane Baugé, Stephanie Bourdon, Benoit Lepetit, and Laurence Pillard for their technical support and help with data collection.

## Author Contributions

CM and PETD conceptualized the research question, collected experimental data and wrote the paper. CV conceptualized the research question, conducted the analyses and wrote the paper. AM, AJ, and KC conceptualized the research question and collected experimental data. MT contributed to conceptualizing the research question. All authors approved the final version of the manuscript for submission.

## Preregistration

This study was preregistered (NCT04593316).

## Data accessibility

The data and materials are currently private for peer review.

---

**Supplementary Fig. 1.**
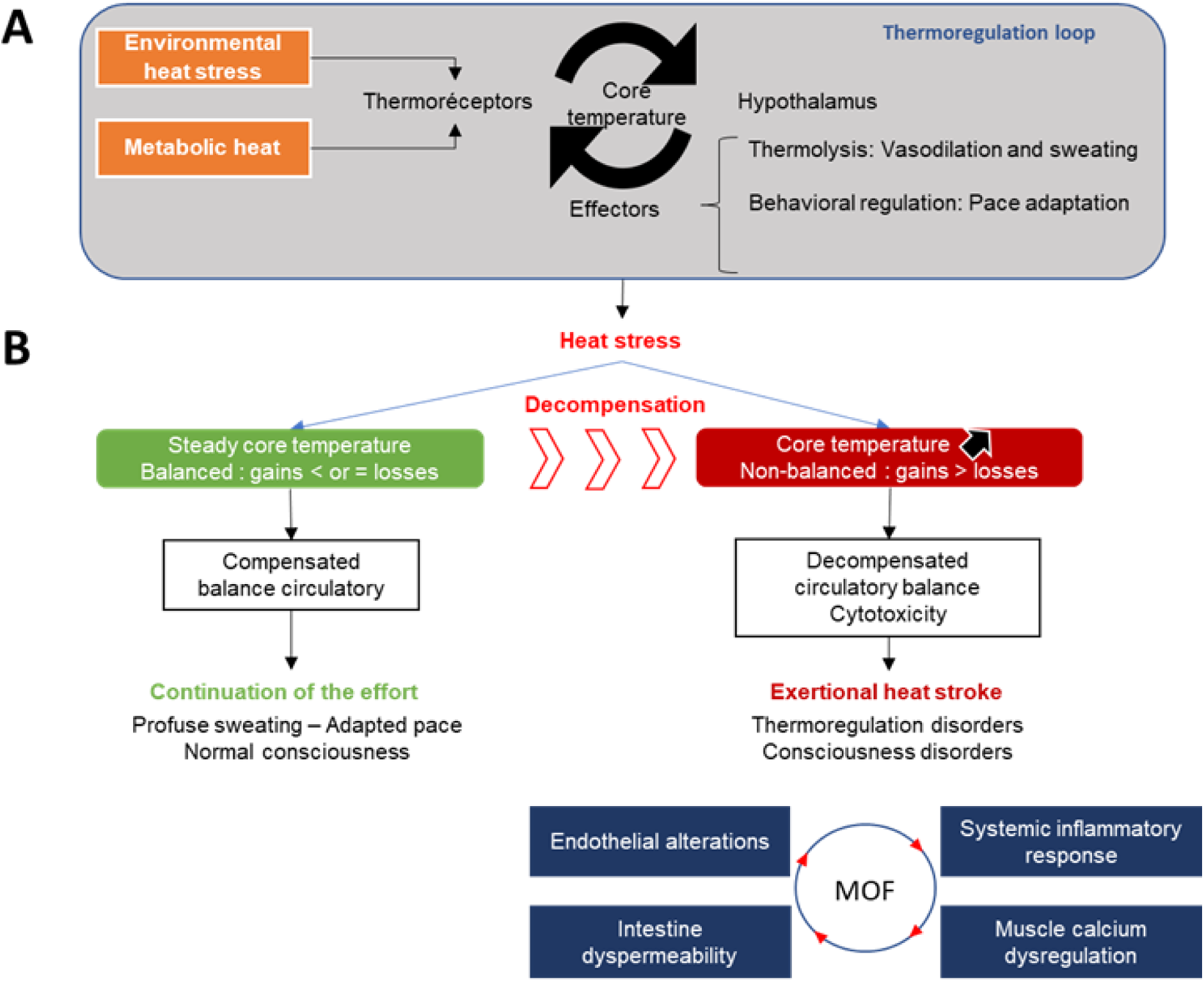
Pathogenesis of exertional heatstroke: a graphical overview of suspected physiological mechanisms. **(A) Physiological thermoregulation.** Whatever exogene (outdoor) and/or endogene (exercise metabolism), heat exposure draws a regulatory loop. Thermoreceptors inform the thermoregulatory centers (hypothalamus) that will coordinate the response of the effectors. Thus, heat dissipation is obtained by generalized subcutaneous vasodilation and sweat evaporation. The reduction of heat stress can also come from behavioral regulation including the adaptation of the pace (decrease in metabolic production) and the search for shelter and the adaptation of the clothing (decrease of the external heat load). **(B) Pathophysiology of exertional heat stroke.** The occurrence of exercise heat stroke proceeds from the transition from a compensation of heat load to a non-compensable heat stress phasis (gains greater than heat losses) occurring when the cardiac output no longer allows to provide for thermoregulation needs. This uncontrolled hyperthermia leads to cytotoxic effects and a systemic inflammatory response that can lead to multi-organ failure. In this context of decompensated circulatory balance, the pathophysiological mechanisms would be based on disorders of intestinal permeability with release of activating molecules of the inflammatory and immune system. Endothelial alterations would also be responsible for coagulopathy. The direct cytotoxic effect of the temperature increase could also induce brain alterations, particularly hypothalamic. In a certain case, mutations in the ryanodine receptor RyR 1 (a muscle receptor involved in the release of intracellular calcium during contraction) could promote the occurrence of this decompensation of thermoregulation to exercise, by the disorders of the excitation coupling contraction that they induce (Epstein and Yanovich 2019, Laitano, Leon et al. 2019).

## Supplementary Methods

### Power considerations

To ensure that our analyses were nevertheless adequately powered, we conducted post hoc power analyses on significant data. For this purpose, we used the GPOWER software with the following parameters: Tails: Two; 𝛼 error probability: 0.05; Total sample size: 94 (51 cases + 43 controls). The effect size Cohen’s d effect size was computed from the reported rank-biserial correlation from each Mann–Whitney U test as follows (Equation 1):

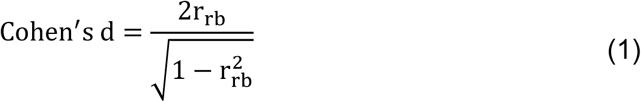

**Supplementary Table 1.** A descriptive and approximate classification scheme for the interpretation of the log scale of Bayes factor BF_10_ (adapted from (Jeffreys 1961)).

| | Log ($BF_{10}$) | Interpretation | Symbol |
| --- | --- | --- | --- |
| Growing evidence in<br>favour of $H_1$ | $> 2$ | extreme evidence for $H_1$ | $H_1^{****}$ |
| | $[1.48 ; 2]$ | very strong evidence for $H_1$ | $H_1^{***}$ |
| | $[1 ; 1.48]$ | strong evidence for $H_1$ | $H_1^{**}$ |
| | $[0.48 ; 1]$ | moderate evidence for $H_1$ | $H_1^*$ |
| | $[0 ; 0.48]$ | anecdotal evidence for $H_1$ | ns |
|  | 0 | no evidence | ns |
| Growing evidence in<br>favour of $H_0$ | $[-0.48 ; 0]$ | anecdotal evidence for $H_0$ | ns |
| | $[-1 ; -0.48]$ | moderate evidence for $H_0$ | $H_0^*$ |
| | $[-1.48 ; -1]$ | strong evidence for $H_0$ | $H_0^{**}$ |
| | $[-2 ; -1.48]$ | very strong evidence for $H_0$ | $H_0^{***}$ |
| | $< -2$ | extreme evidence for $H_0$ | $H_0^{****}$ |

Log( $BF_{10}$ ): log scale of Bayes factor $BF_{10}$ ; $H_1$ : alternative hypothesis; ns: non-significant; $H_0$ : null hypothesis

**Supplementary Table 2.** Descriptive statistics for the Multidimensional Assessment of Interoceptive Awareness (MAIA) questionnaire in cases with a history of exertional heatstroke and controls.

|  | **Cases**<br>**(n=51)** |  | **Controls**<br>**(n=43)** |  |
| --- | --- | --- | --- | --- |
|  | M | SD | M | SD |
| Scale **Noticing** | 3.52 | 0.87 | 3.88 | 0.72 |
| Scale **Not-distracting** | 2.46 | 0.89 | 2.77 | 0.90 |
| Scale **Not-worrying** | 2.91 | 0.89 | 3.02 | 0.93 |
| Scale **Attention regulation** | 3.05 | 0.89 | 3.55 | 0.76 |
| Scale **Emotional awareness** | 3.44 | 0.95 | 3.88 | 0.81 |
| Scale **Self-regulation** | 2.97 | 1.15 | 3.50 | 0.98 |
| Scale **Body listening** | 2.37 | 1.10 | 3.05 | 1.10 |
| Scale **Trusting** | 3.80 | 0.89 | 4.11 | 0.96 |
| **Total score** | 24.53 | 4.81 | 27.76 | 4.52 |

M: mean; SD: standard deviation

**Supplementary Table 3.** Descriptive statistics for the Global Motivation Scale (GMS) questionnaire in cases with a history of exertional heatstroke and controls.

|  | **Cases**<br>**(n=51)** |  | **Controls**<br>**(n=43)** |  |
| --- | --- | --- | --- | --- |
|  | M | SD | M | SD |
| Scale ***IM to know*** | 21.61 | 4.51 | 22.54 | 3.89 |
| Scale ***IM to accomplishment*** | 22.25 | 4.15 | 22.07 | 4.73 |
| Scale ***IM to stimulation*** | 20.47 | 4.70 | 20.56 | 5.04 |
| Scale ***Identified regulation*** | 21.92 | 3.74 | 21.58 | 3.87 |
| Scale ***Introjected regulation*** | 18.18 | 5.10 | 17.93 | 5.54 |
| Scale ***External regulation*** | 18.96 | 5.23 | 17.33 | 5.96 |
| Scale ***Amotivation*** | 11.43 | 4.08 | 14.51 | 5.59 |

M: mean; SD: standard deviation; IM: intrinsic motivation

**Supplementary Table 4.**
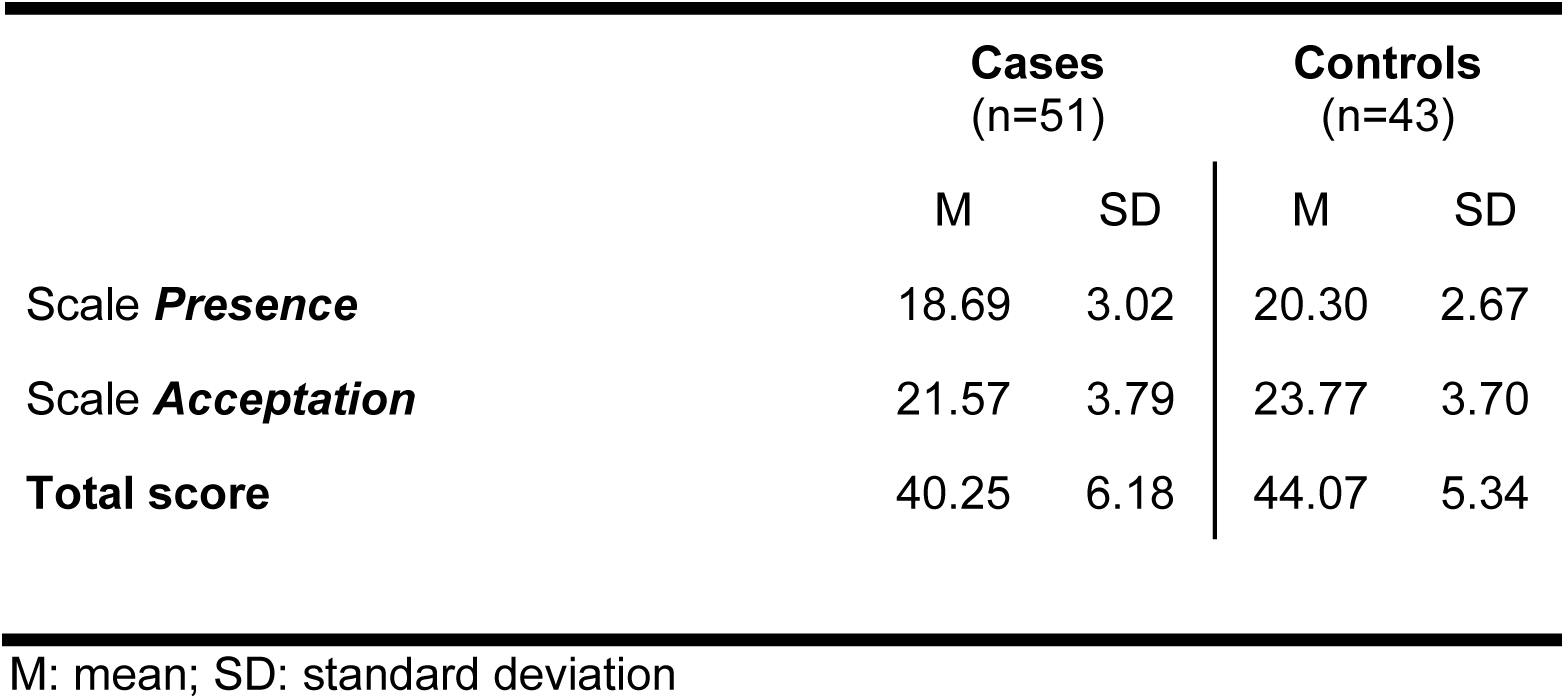
Descriptive statistics for the Freiburg Mindfulness Inventory in cases with a history of exertional heatstroke and controls.

|  | **Cases**<br>(n=51) |  | **Controls**<br>(n=43) |  |
| --- | --- | --- | --- | --- |
|  | M | SD | M | SD |
| Scale ***Presence*** | 18.69 | 3.02 | 20.30 | 2.67 |
| Scale ***Acceptation*** | 21.57 | 3.79 | 23.77 | 3.70 |
| **Total score** | 40.25 | 6.18 | 44.07 | 5.34 |

M: mean; SD: standard deviation

**Supplementary Table 5.** Post hoc power analysis for interoceptive and motivation dimensions showing significant group differences.

| | rb<br>(midpoint of CI) | Cohen's d | Achieved Power (1-β) |
| --- | --- | --- | --- |
| Scale ***Noticing*** | 0.22 | 0.45 | 0.79 |
| Scale ***Attention regulation*** | 0.32 | 0.68 | 0.93 |
| Scale ***Emotional awareness*** | 0.29 | 0.60 | 0.85 |
| Scale ***Self-regulation*** | 0.28 | 0.57 | 0.82 |
| Scale ***Body listening*** | 0.33 | 0.70 | 0.94 |
| Scale ***Amotivation*** | 0.32 | 0.68 | 0.93 |

Rbs: rank-biserial correlation; CI: confidence interval;

